# Mapping under-five mortality in Malawi: towards achieving sustainable development goals

**DOI:** 10.1101/2024.03.11.24304141

**Authors:** Ruth Vellemu, Melody Sakala, Evetta Kuwala Chisope, James Chirombo

## Abstract

As many low- and middle-income countries continue making gains towards attaining sustainable development goals in under-five mortality, surveillance of mortality outcomes and indicators at the sub-district level will become important as national- and district-level estimates may mask areas with a high burden. Spatial statistical modelling techniques such as geostatistical prevalence mapping can play a role in identifying hotspots of relatively high under-five mortality. To achieve this objective, it is necessary to combine data from multiple sources with different spatial resolutions to produce maps that reveal these hotspot clusters. We pooled DHS, high-resolution census data, economic vulnerability, and malaria risk data to estimate under-five mortality at a sub-district level in Malawi. Using a Bayesian hierarchical modelling approach, we fitted a binomial generalized linear geostatistical model with local area effects to generate estimates of under-five mortality at a sub-district level in Malawi. Results, in general, showed low mortality rates across the country with pockets of locations mostly in northern Malawi showing elevated risk. Rural locations were associated with higher odds of under-five mortality compared to urban locations. Our study provides a better understanding of progress made after the Millennium Development Goals in 2015 and can help improve surveillance through the application of targeted interventions which can lead to the attainment of sustainable development goals by 2030.

## Introduction

Efforts to improve child health and well-being have accelerated in recent years, first with the adoption of Millennium Development Goals (MDGs) [1] until 2015 and after that the Sustainable Development Goals (SDGs) [2]. Globally, under-five mortality rate (U5MR) reduced from 93 deaths per 1000 live births in 1990 to 37.7 deaths per 1000 live births in 2019, representing a 59% decline [3,4]. However, several low and middle-income (LMIC) countries especially in Sub-Saharan Africa (SSA) did not achieve the MDG 4 target of reducing U5MR between 1990 and 2015 by two-thirds with only 11 out of 48 countries meeting the target [5]. In the SDG era, the target is for every country to achieve U5MR of not more than 25 per 1000 live births by 2030.

Despite the failures in some SSA settings to meet MDG 4, notable progress has been made. For example, Malawi is among the 11 countries in Africa to have achieved the MDG 4 target, having reduced U5MR from 247 deaths per 1000 in 1990 to 73 deaths per 1000 by 2013 [6]. Several interventions have been credited for this achievement in Malawi, including the adoption of integrated community case management (ICCM) for childhood diseases in 2007 [7,8]. ICCM has been a critical means for treating common illnesses such as diarrhoea, pneumonia and malaria resulting in reduced mortality through improved coverage and access for hard-to-reach populations [7–9].

For Malawi to achieve the SGD target of reducing U5MR to 25 per 1000 live births by 2030, the gains that have been made until now must be sustained and improved upon. There is a need to adapt interventions to address the emerging challenges. For example, novel pandemics such as COVID-19 can stretch government resources from highly effective programmes. A recent modelling study across 18 countries including Malawi, found that disruptions to healthcare service utilizations had an impact on child mortality [10]. These disruptions led to an average 3.6% increase in mortality across the 18 countries. In Malawi, there was a 4.3% relative increase in under-five mortality [10]. Other disruptions such as health financing, HIV, and TB services also contributed to increased child mortality [11].

Apart from pandemic threats, heterogeneities in mortality persist despite a sustained downward trend resulting in some sections of the population bearing higher burdens, posing challenges to attaining SGDs. Similar challenges persist across Africa despite a continuous downward trend in U5MR across many countries. For example, a study on child mortality in Senegal, Rwanda and Uganda found that mortality had decreased in all the 3 countries [12]. Kenya registered an overall impressive decline in U5MR between 1965 and 2013. Yet, some barriers that prevented meeting the MDG 4 targets still exist and if not addressed can potentially derail progress towards achieving SDGs. For example, in Kenya, variations were observed in the reduction in U5MR both between and within counties [13]. These inequalities in mortality levels are possibly due to factors such as differences in deprivation, access to health care and cultural norms. Inequalities have also been found in Senegal, Rwanda, and Uganda [12]. It has been projected that 94.4 million children under 5 years will die if 2015 mortality rates remain constant while 68.8 million will die if each country continues to reduce its mortality at the pace estimated between 2000-15 [4]. With the recent 2020-2022 COVID-19 pandemic, there are concerns that global progress towards achieving SDGs including reducing child mortality will be negatively impacted [14]. Therefore, there is a need to substantially scale up interventions to reduce the inequalities in mortality reduction across most of SSA. Even the countries that achieved MDG targets must remain vigilant to achieve the SDGs by 2030. Malawi faces similar challenges. The heterogeneities observed in countries such as Kenya are observed in Malawi too. Differences in child health indicators between urban-rural areas of Malawi have been well documented with a U5MR of 77 deaths per 1000 live births in rural compared to 60 deaths per 1000 live births in urban areas. Heterogeneities also exist within localized areas [15,16]. For example, within urban areas, there is evidence of heterogeneities in child health outcomes such as under-five mortality driven by poor outcomes among the poor residing in urban slums [16].

To identify areas with elevated U5MR within countries, geospatial approaches are increasingly being used to map the spatial variation in risk and hence provide stakeholders with clear and actionable output and recommendations. Geostatistical approaches use data collected at sparse discrete locations to provide a continuous surface covering the entire area allowing for the estimation of prevalence at unsampled locations [17,18]. Various geostatistical models have been fitted for various diseases and child mortality in Africa [19,20]. Where there is data aggregated at an administrative unit level such as a district, geospatial approaches are also used to estimate and map the relative risk of the outcome. Unlike diseases such as malaria for which there are national surveys such as the malaria indicator survey (MIS) and various community surveys, mortality data in many developing countries typically comes from large periodic surveys such as the demographic and health surveys (DHS). These surveys involve sampling at discrete locations, and it is important to get the maximum value from the data. This makes the use of spatial statistical techniques a necessary undertaking to obtain estimates in unsampled locations.

In this paper, we aimed to model under-five mortality at the sub-district level to reveal the heterogeneities in U5MR reduction efforts that exist in Malawi, to assess progress towards SDGs. We used a binary generalized linear geostatistical model to generate smooth maps to estimate U5MR at unsampled locations throughout Malawi to reveal locations with elevated mortality rates that may be masked by district-level estimates of mortality [21].

## Methods

### Study setting

Malawi is a small landlocked country in Southern Eastern Africa, sharing boundaries with Zambia, Tanzania, and Mozambique. The country is divided into 3 administrative regions and further into 28 districts. The public health system comprises 4 central hospitals, district hospitals and health centers. At the community level, health surveillance assistants (HSAs) provide basic primary care including treatment of common illnesses in children. In addition, the faith-based health facilities primarily under the Christian Health Association of Malawi (CHAM) also provide a wide network of health facilities. Fig S1 in the supplementary materials provides more information about the districts and the distribution of public health facilities in Malawi.

### Mortality data

Data for this study were obtained from the 2015-16 Malawi Demographic and Health Survey (MDHS) [15]. The MDHS is a stratified nationally representative survey that uses a two-stage cluster sampling method and is conducted every five years [15]. DHS surveys are conducted in many countries across the globe, and they provide estimates for a range of indicators thus allowing comparison between countries [22]. At the first stage, clusters (i.e., enumeration areas) are selected using a probability sampling proportion to size approach. The second stage involves the selection of households from which eligible participants are interviewed. The 2015-16 MDHS enrolled 24,562 women respondents aged between 15-49 years [15]

### Census and population level covariates data

In addition to the point-level covariate data from the MDHS referenced at each cluster, we also obtained population-level geospatial datasets covering the whole country from Humanitarian Data Exchange. We used vulnerability scores to capture the social-economic structure of the country. Additionally, being a malaria-endemic country and given its status as the leading cause of mortality in children, we included malaria risk as a potential predictor of mortality [23]. The inclusion of these covariates was influenced by studies that have shown that low socio-economic status and malaria are among the factors contributing to child mortality [24]. Lastly, population data at 100 m spatial resolution were extracted from the WorldPop data sets. These data provided estimates at a high resolution thus allowing the calculation of important health indicators at a finer scale than would be possible if only district-level covariates were used.

### Dependent and independent variables

The outcome variable was the survival status of a child categorized as ‘yes’ for alive and ‘no’ for dead. Individual level variables from DHS included mothers’ age, mother’s level of education categorized as primary and below and secondary and above, area of residence classified as rural and urban and child’s birth weight. We also extracted malaria risk and vulnerability, both captured as percentages at each cluster location (Supplementary material, Fig S2).

### Ethical approval

The study used data from the 2015/16 MDHS survey. Access to the data was granted by the DHS program upon registration. Ethical approval for the MDHS was obtained from the National Health Sciences Research Committee (NHSRC). A written consent was obtained during the survey before any interview. For children and adolescent’s participation, consent was provided by their parents/guardians. No individual identifiers were used in this secondary analysis.

### Data analysis

Descriptive statistics were used to summarize study variables. We generated a table of baseline characteristics to understand key socio-demographic properties. Exploratory spatial mapping of cluster distribution within districts in Malawi and the underlying population densities was also done. To investigate factors affecting under-five mortality, all potential variables were fitted into a geostatistical model. All data analyses were done in the R environment for statistical computing using the geostatsp package [25] which uses the integrated nested Laplace approximation (INLA) on the backend [26].

### Statistical modelling

Let *n*(*x*_*i*_) be the number of children that died and *N*(*x*_*i*_) be the total number of births recorded at location *x*_*i*_ respectively. We assumed that the number of deaths follows an independent binomial distribution with the number of trials *N*_*i*_ and probability of a positive outcome *p*(*x*_*i*_). The probability of death can then be modelled as a binomial generalised linear geostatistical model (GLGM) as follows.

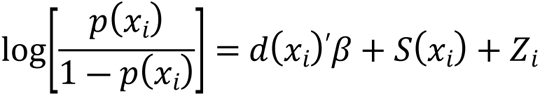

The term *d*(*x*_*i*_)^′^ contains both individual level covariates measures such as age of the women and population level covariates such as malaria risk and vulnerability score of the location. For the spatially correlated term *S*(*x*_*i*_), a Markov random field approximation of the Matern correlation function on a regular lattice was used. The term *Z*_*i*_ captures independent random effects. For the spatially structured random effects, we assumed an isotropic and stationary spatial process with an exponential correlation function [25]. For our analysis, model building was based on the known contribution of the variables to mortality based on literature. Variables were thus added to the model based on their clinical significance. We did not fit univariate models to test the significance of a variable before adding it into the model. Such an approach has the unintended consequence of omitting variables that are important in predicting the outcome variable.

## Results

### Study participants

Table 1**Error! Reference source not found.** shows the baseline characteristics of the study participants. There were a reported 286 deaths which was 2.5% of the recorded births. These deaths were roughly equally distributed between urban and rural (2.2% and 2.5%) respectively. There was almost equal distribution of deaths between male and female children (2.2% and 2.7% respectively). In general, the proportions of the key demographic characteristics between the outcome categories were comparable.

**Table 1:**
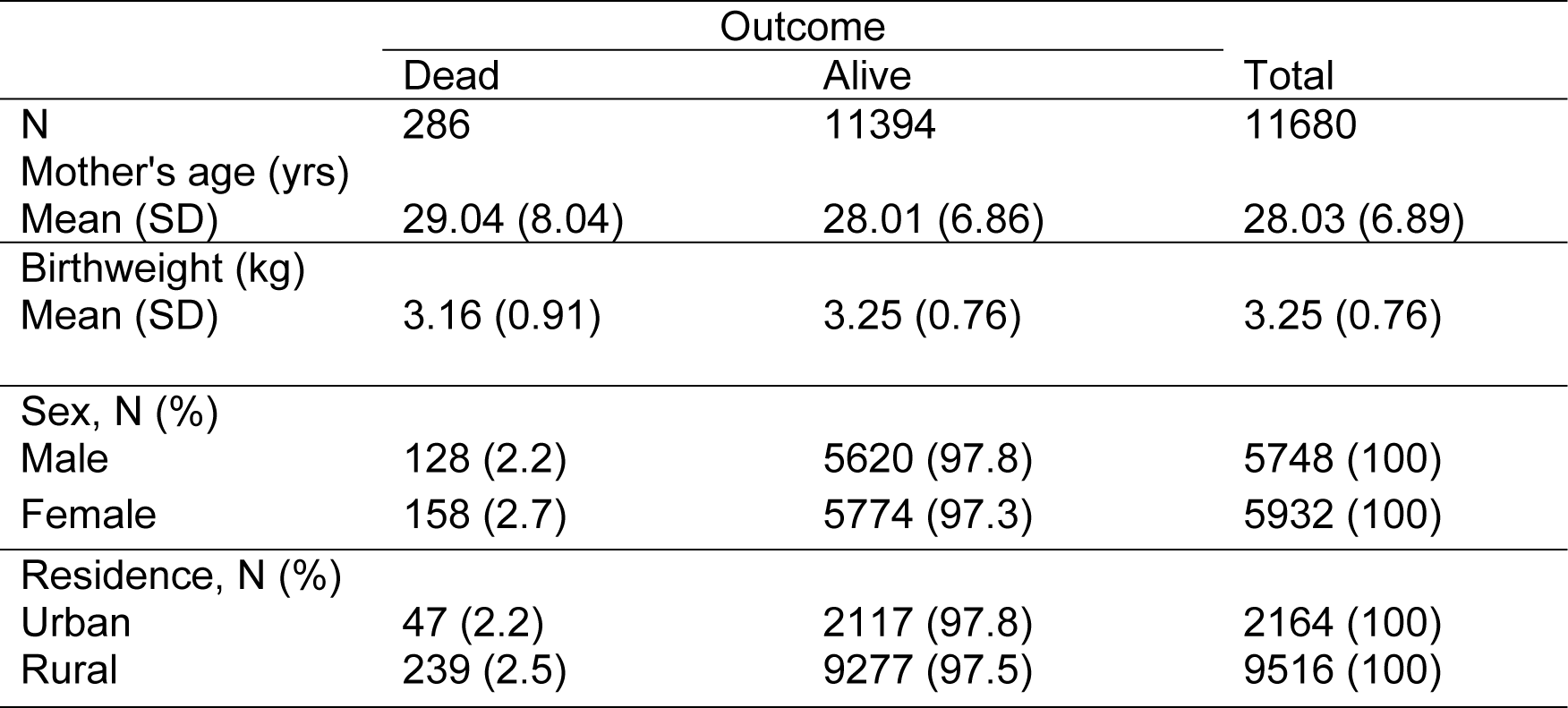
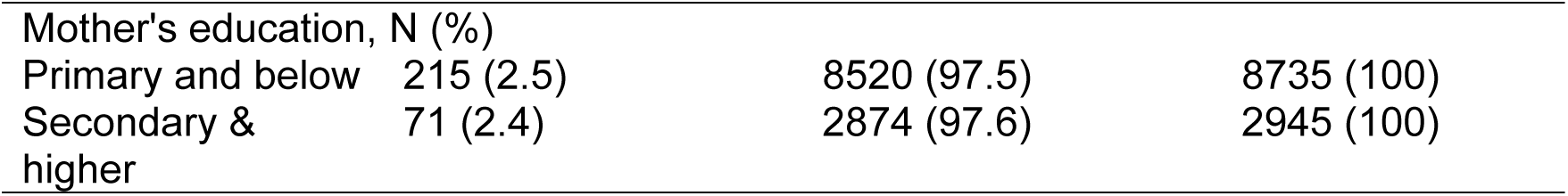
Baseline characteristics of mothers with children under the age of five in Malawi averaged across all clusters.

### Cluster locations and population density

Fig 2 shows cluster locations within districts from which respondents for the survey were obtained and the underlying population density. High population density is observed in the cities of Mzuzu, Lilongwe and Blantyre. Lakeshore areas also have relatively higher population density.

**Fig 1:**
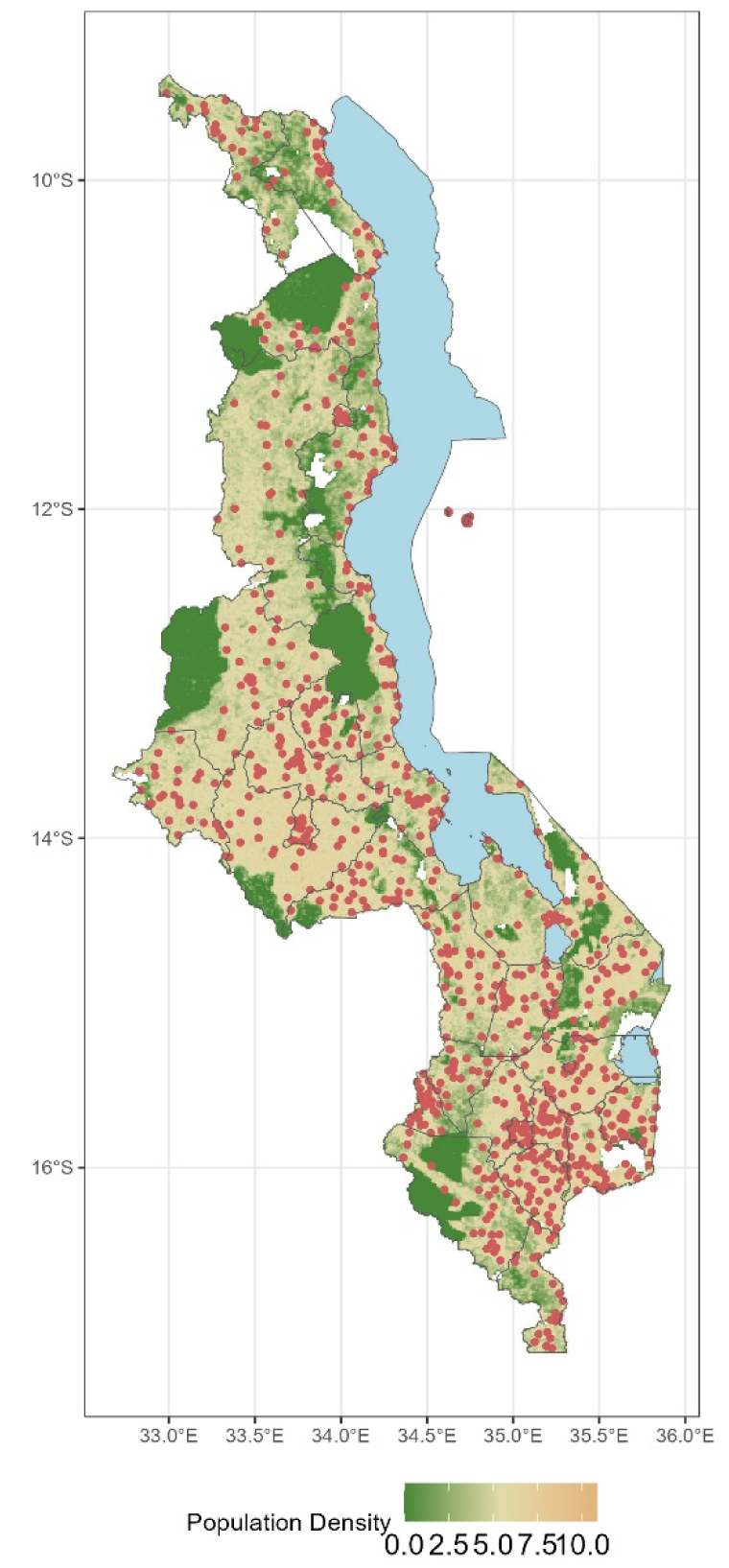
Cluster locations and the underlying population density on the log scale. The green spots are protected areas such as national parks and forest reserves.

### Under-five mortality rates

Fig 2 shows crude under-five mortality rates at the district level. It is observed that districts with the highest under-five mortality rate are mostly in the northern and southern parts of Malawi followed by the central region. However, all three cities namely Mzuzu, Lilongwe and Blantyre from northern, central, and southern regions respectively have lower rates of under-five mortality.

**Fig 2:**
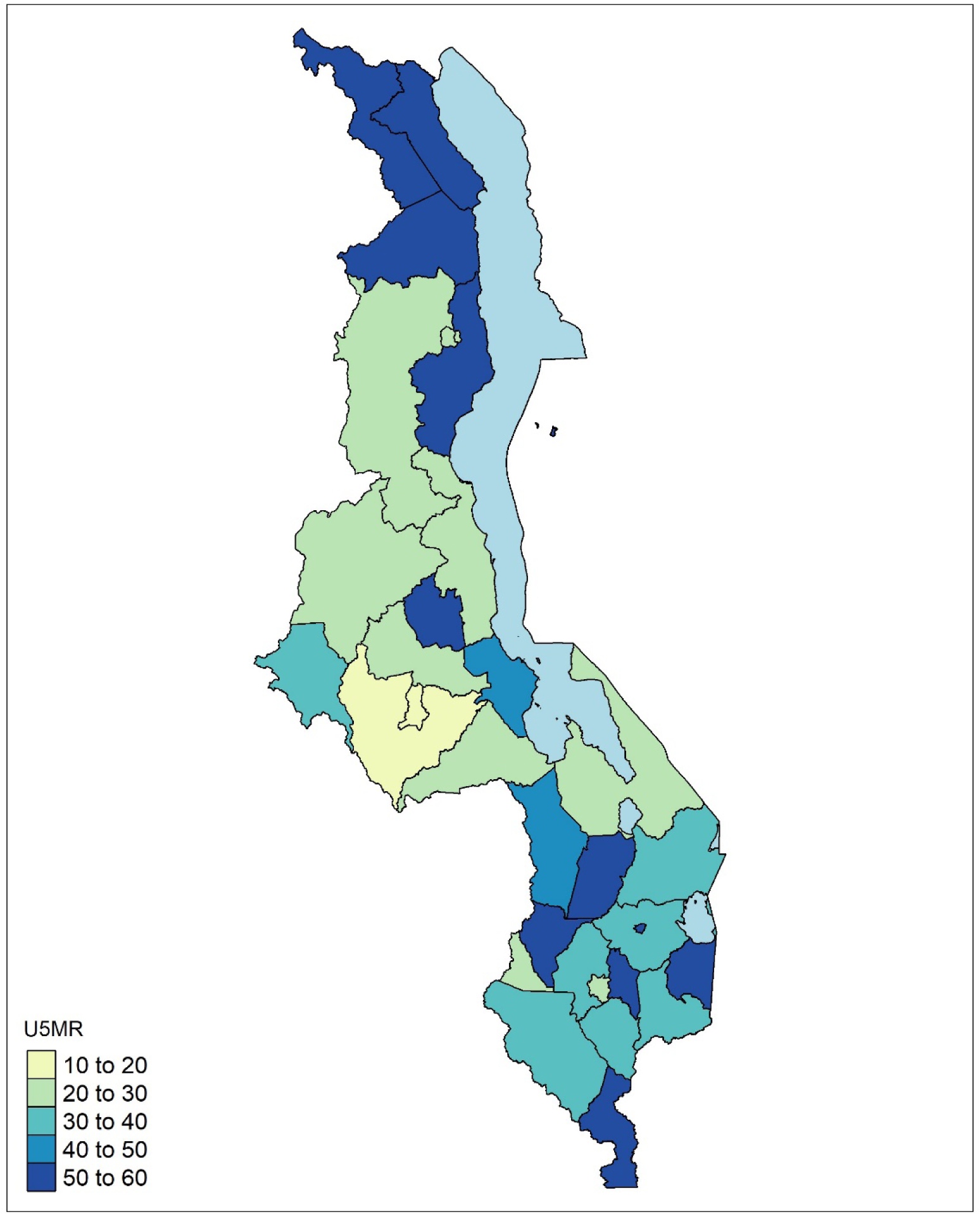
Aggregate child mortality estimates at the district level. Mortality ranges from less than 20 deaths per 1000 to over 50 deaths per 1000 live births. The lowest mortality is observed in the 3 main cities of Mzuzu, Lilongwe and Blantyre

### Model results

Table 2 presents posterior estimates from the geostatistical model. The effect of mother’s age was very marginal. For a unit increase in age, there was only a 1% increase in the odds of mortality (OR=1.01, 95% CI=0.99-1.02). The effect of birthweight was neutral (OR=1.00, 95% CI=1.00-1.02).

**Table 2:** Posterior estimates of the binomial geostatistical regression model.

|  | OR | 2.5% quintile | 97.5% quintile |
| --- | --- | --- | --- |
| Mother's age | 1.01 | 0.99 | 1.02 |
| Birthweight | 1.00 | 0.98 | 1.01 |
| Residence |  |  |  |
| Urban | REF |  |  |
| Rural | 1.22 | 1.12 | 1.33 |
| Mother's education |  |  |  |
| Secondary and above | REF |  |  |
| Primary and below | 1.04 | 0.91 | 1.18 |
| Vulnerability | 1.00 | 1.00 | 1.02 |
| Malaria risk | 1.00 | 0.99 | 1.03 |

Children residing in rural areas had a 22% increase in the odds of mortality compared to their counterparts in urban areas (OR= 1.22, 95% CI=1.12-1.33). Malaria risk was not found to be associated with under-five mortality (OR=1.00, 95% CI=0.99-1.03). Likewise, vulnerability was not found to be associated with mortality (OR = 1.00, 95% CI = 1.00-1.02).

### Predicted mortality risk

Figure 3 shows the predicted mortality risk map. Overall, the map shows heterogeneities in under-five mortality risk. The northern areas in general have a higher risk of mortality especially in Rumphi, Mzimba and parts of Nkhata Bay districts. Lower risks are observed in central and southern areas. The map shows varying risks within districts identifying the within-district heterogeneities that exist.

**Figure 3:**
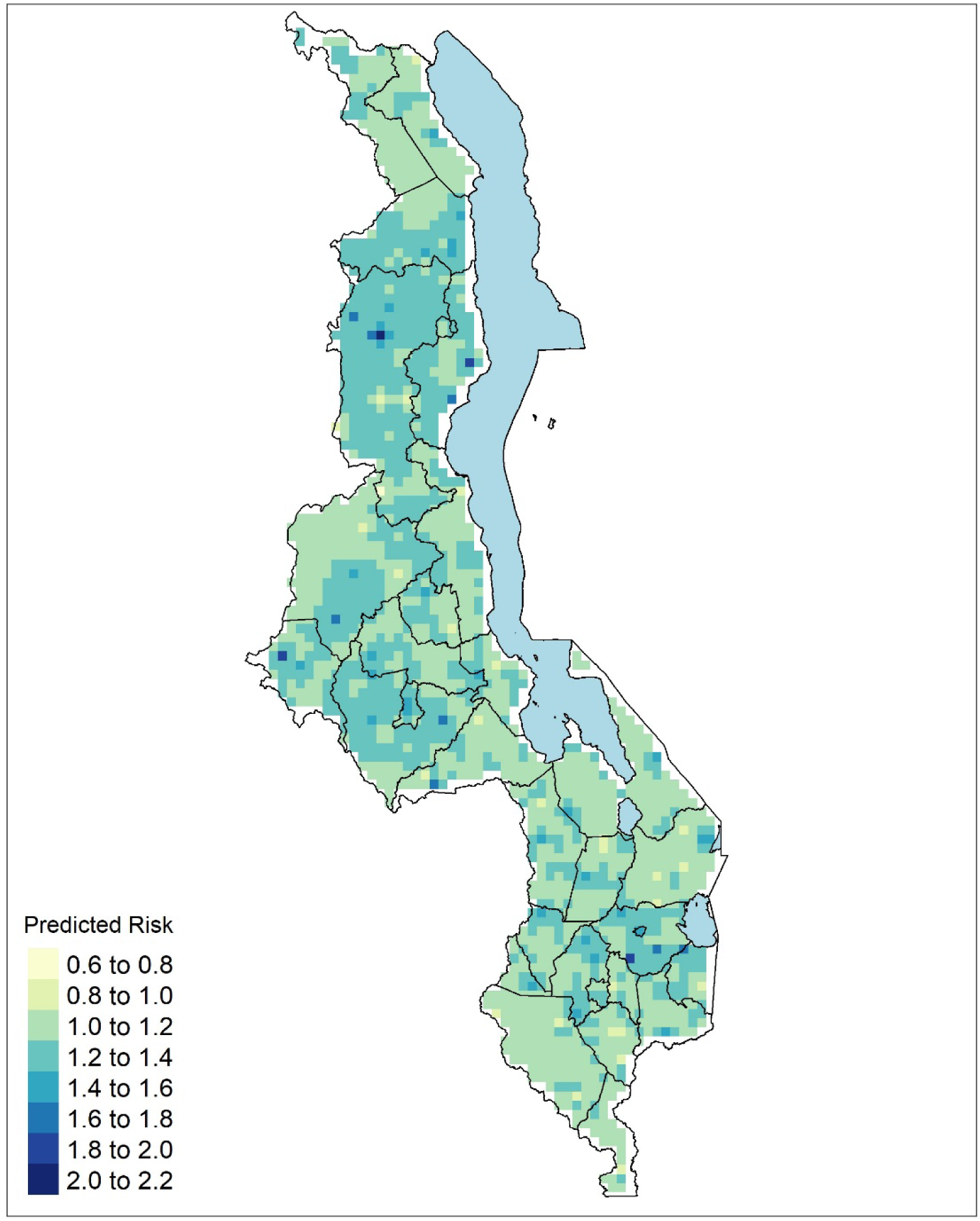
Predicted relative risk estimates of child mortality expressed as a percentage. Dark green zones show areas of elevated risk. Pale zones are areas with the lowest risk.

## Discussion

In this study, we mapped the under-five mortality across Malawi to show spatial heterogeneities that should be taken into consideration in the continued efforts to reduce child mortality to meet the SDGs by the year 2030. There are still pockets of locations with relatively high under-five mortality rate that needs focused interventions to achieve the SDG goal of reducing under-five mortality to less than 25 deaths per 1000 live births. In this study, we looked at the various factors that are still important potential drivers of child mortality in Malawi that should be addressed to help sustain the downward trend and hence achieve SGDs. Our study showed that under-five mortality was associated with the area of residence. This finding agrees with past studies in Malawi that have found that residence is positively associated with under-five mortality. In particular, the risk of children dying was higher for rural respondents compared to their urban counterparts [27,28]. Similar results showing rural-urban differential have been found in other developing countries. For example, in India, under-five mortality was 4.62% in rural areas compared to 3.28% in urban areas [29]. A rural-urban gap was also found in 35 countries in Sub-Saharan Africa [30].

The higher risk of mortality in rural areas compared to urban locations could be because of a lack of adequate healthcare and not following proper disease prevention strategies attributed to poor settings. Such poor setting factors would include poor healthcare-seeking behaviour in rural areas [31], not sleeping under bed nets to prevent malaria, and malnourishment resulting from a shortage of food and balanced meals. Rural areas also generally have a sparse network of health facilities leading to longer travel times to access healthcare which has been shown to lead to a greater risk of mortality [32]. It is believed the urban/rural mortality differentials are attributed to various socio-economic differences that exist within the country. In addition, factors such as better education, more public infrastructure that provides sanitation services, safer water supply, better systems to handle household waste and excreta removal, and easier access to healthcare services that are more favourable in urban than in rural areas can also explain this relationship [33]. The lack of association between under-five mortality and variables such as education, malaria, vulnerability, and mother’s age could be partly attributed to its general decline across the board. This result helps confirm the observations that heterogeneities exist at the subdistrict level which may not be well-captured by district-averaged covariates. Although these variables did not individually show any association with mortality in our study, differences in education, and mother’s age between rural and urban likely contribute to the observed differences in rural-urban under-five mortality [34,35].

Our predicted risk maps indicate differences in mortality within the districts that could be masked by district-level estimation. Many factors could be responsible for relatively higher risk areas in some parts of the country including the role of infectious diseases such as HIV/AIDS and malaria [36,37]. Malaria is endemic to Malawi and has also been shown to be a key risk factor for child mortality [23,28]. Despite not being statistically significant in the model, the high malaria prevalence among children likely substantially contributes to under-five mortality. For example, the 2017 Malawi Malaria Indicator Survey (MIS) reported a prevalence rate of 24% [38]. The varying quality of health systems could also be a driving force behind the observed heterogeneities in under-five mortality within districts.

Our study has a few limitations. Firstly, we used data from the 2015/16 DHS. Therefore, the data are not very recent. DHS surveys are typically done every five years. However, the comprehensive nature of DHS surveys makes them quite useful for the period between successive surveys. It should also be noted that the high-resolution population estimates from WorldPop are modelled estimates and not observed population values. Modelling approaches are used to estimate population density at a remarkably high resolution from national census data. Furthermore, recall bias is another problem encountered in DHS surveys when historical information is sought. Another limitation of this study is due to potential confounding which can affect the reported associations. As this was an observation study, we did not actively control for all known confounding factors.

## Conclusion

Our results showed that inequalities in the reduction of under-five mortality exist within districts in Malawi despite the general decline in under-five mortality across the country. There are areas where the risk is higher than the national average. Our modelling approach provides a picture of the heterogeneities at the sub-district level. Investigating these sub-national inequalities provides policymakers with valuable information on the state of child mortality and will help shape district-specific efforts towards achieving SDGs. To further reduce under-five mortality, efforts to close the gap in healthcare utilization, distribution of resources and the uneven use of interventions between rural and urban locations must be intensified.

## Data Availability

The data can be accessed upon request from the DHS program on https://dhsprogram.com/

## Acknowledgements

RV thanks the DELTAS Africa Initiative, Sub-Saharan Africa Consortium for Advanced Biostatistics Training (SSACABT) for scholarship to pursue a MSc at the University of Malawi. We also thank the DHS program for granting free access to the 2015/16 Malawi DHS data.

## Notes

### Competing Interest Statement

The authors have declared no competing interest.

### Author Declarations

The DHS was approved by the National Health Sciences Research Committee

